# A follow-up study of children infected with SARS-CoV-2 from Western China

**DOI:** 10.1101/2020.04.20.20073288

**Authors:** Hongmei Xu, Enmei Liu, Jun Xie, Rosalind Smyth, Qi Zhou, Ruiqiu Zhao, Na Zang, Xiaoru Long, Yuyi Tang, Janne Estill, Shu Yang, Jing Zhu, Xiaofeng Yan, Fang Gong, Wenguang Tian, Xiaqia Zhou, Yunbo Mo, Hongzhou Xiao, Zhengzhen Tang, Yanni Chen, Yi Wang, Yuxia Cui, Xiuling Fang, Feiyu Li, Yong Tian, Peibo Li, Quanmin Deng, Chongsong Ren, Ronghui He, Yi Li, Hong Qin, Aiguo Wang, Hongli Deng, Jianguo Wu, Wenbo Meng, Weiguo Li, Yao Zhao, Zhengxiu Luo, Zijun Wang, Yaolong Chen, Gary Wong, Qiu Li

## Abstract

**Background:** To clarify the characteristic and the duration of positive nucleic acid in children infected with severe acute respiratory syndrome coronavirus 2 (SARS-CoV-2), including asymptomatic children.

**Methods:** A total of 32 children confirmed with SARS-CoV-2 infection between January 24 and February 12, 2020 from four provinces in Western China were enrolled in this study and followed up until discharge and quarantine 14 days later.

**Results:** Eleven children (34%) were asymptomatic, among whom six children had normal computed tomographic (CT) scan images. Age and gender were not associated with clinical symptoms or the results of CT scan in children infected with SARS-CoV-2. The concentrations of white blood cells and neutrophils were higher in children with asymptomatic infection than in children with clinical symptoms or CT abnormalities. Patients who presented with CT abnormalities had lower D-dimer or lower total bilirubin than those who had normal CT scan but clinical symptoms. All children recovered and no one died or was admitted to the pediatric intensive care unit (PICU). The mean duration of positive SARS-CoV-2 nucleic acid was 15.4 (SD=7.2) days and similar for both asymptomatic children and children with symptoms or CT abnormalities. We found a significant negative correlation between the lymphocyte count and the duration of positive nucleic acid test.

**Conclusions:** Children with asymptomatic infection should be quarantined for the same duration as symptomatic patients infected with SARS-CoV-2. The clinical significance and mechanism behind the negative correlation between the number of lymphocytes and the duration of positive SARS-CoV-2 needs further study.

## 1. BACKGROUND

SARS-CoV-2 was first detected when a cluster of patients with pneumonia of unknown cause emerged in Wuhan, China in December, 2019. On February 11, the World Health Organization (WHO) officially named the disease as “Corona Virus Disease hyphen one nine” (COVID-19) ^[1-6]^. As the number of infections and deaths continued to rise, WHO declared COVID-19 as a global pandemic on March 11, 2020 ^[7]^. and is causing serious health and economic impact in 179 countries and regions worldwide. COVID-19, together with severe acute respiratory syndrome (SARS) and Middle East respiratory syndrome (MERS), is a disease caused by beta-coronavirus. However, the mortality rate of COVID-19 is lower than that of SARS (more than 40% in people over 60 years old) and MERS (30%) ^[8-11]^.

At present, many papers have reported on the epidemiology and clinical characteristics of COVID-19. However, most studies have so far focused on adults ^[12-14]^ and several case series of children have been limited by small sample size or the lack of follow-up, in particular for asymptomatic patients ^[15-20]^. Children, compared with adults, have a developing immune system and different physical characteristics, thus it is not appropriate to directly use evidence from adults to guide clinical practice of COVID-19 in children. Therefore, this is the first follow-up study that included 32 children infected with SARS-CoV-2 from multiple centers in four provinces of western China, to clarify the clinical characteristics, the duration of positive nucleic acid tests and their correlation with lymphocyte counts between children with asymptomatic infection and those with clinical symptoms, thus providing evidence to guide management of children infected with SARS-CoV-2.

## 2. Method

### 2.1 Study design and patients

This multicenter retrospective study was approved by the Ethics Committee of Children’s Hospital of Chongqing Medical University (No. 2020-002). Given the urgency in policy and clinical decision-making for COVID-19 and difficulty of confirming information for most patients, informed consent was exempted for all patients after discussion and approval of the above-mentioned committee. We recruited 32 children (aged less than 18 years) with confirmed SARS-CoV-2 infection between January 24 and February 12, 2020 in several local hospitals designated for the diagnosis and treatment of COVID-19 from Chongqing Municipality, Guizhou Province, Shaanxi Province and Sichuan Province. All cases were diagnosed based on the WHO interim guidance ^[21]^. And all the cases were followed up two weeks later after discharge in the local hospital.

### 2.2 RT-PCR detection

Specimens of throat swab were obtained and sent to the Chinese Centers for Disease Control and Prevention (CDC) for testing in a unified and standardized way of transportation. RNA was extracted and tested by real-time RT-PCR with SARS-CoV- 2 specific primers and probes. The Chinese Center for Disease Control and Prevention recommends: RT-PCR targeting open reading frame (ORF1ab), primers and probes of nucleoprotein (N) gene region to test for SARS-CoV-2. Two genes were targeted during the real-time RT-PCR assay : Target 1 (ORF1ab): forward primer (F) CCCTGTGGGTTTTACACTTAA, reverse primer (R) ACGATTGTGCATCAGCTGA, and fluorescent probe (P) 5’-FAM-CCGTCTGCGGTATGTGGAAAGGTTATGG-BHQ1-3’; and Target 2 (N): forward primer (F): GGGGAACTTCTCCTGCTAGAAT, reverse primer (R) CAGACATTTTGCTCTCAAGCTG, and fluorescent probe (P) 5’-FAM-TTGCTGCTGCTTGACAGATT-TAMRA-3’. The results were classified into three categories: a cycle threshold value (Ct-value) less than 37 was defined as a positive test result, and a Ct-value of 40 or more was defined as a negative test. A medium load, defined as a Ct-value of 37 to less than 40, required confirmation by retesting. If the repeated Ct-value was less than 40 and an obvious peak was observed, or if the repeated Ct-value was less than 37, the retest was deemed positive ^[22]^.

### 2.3 Data collection

We obtained the electronic medical records of the confirmed COVID-19 children from the Hospital Information System (HIS) and Laboratory Information System (LIS) of each designated hospital for diagnosis and treatment of COVID-19. We extracted the following five types of information: 1) history of exposure: history of traveling and close contacts with areas heavily affected by COVID-19 (Wuhan), history of exposure to the Huanan seafood market, or family cluster of the disease; 2) demographic characteristics: age, gender, vital signs, weight, height, and comorbidities; 3) laboratory findings: routine blood test results, blood biochemistry, coagulation function, infection markers and detection results of common respiratory pathogens; 4) radiological findings: chest X-ray and CT scans; and 5) clinical outcomes: Recovery was defined as the patient being discharged after having normal body temperature for more than three days, no respiratory symptoms, CT scan changes resolved, and two consecutive negative test results of SARS-CoV-2 (with at least one day between tests). Asymptomatic infection was defined as the patient being tested positive for SARS-CoV-2 without manifesting any clinical symptoms or abnormal chest imaging findings. Severe cases were defined as having at least one of the following: 1) increased respiratory rate, dyspnea, or cyanosis of the lips; 2) pulse oxygen saturation ≤93%, or oxygen concentration ≤ 300 mmHg in arterial partial pressure inspiration, or partial pressure of oxygen in arterial ≤ 300 mmHg in inspiration; or 3) lobular lesion or lesion with a progression to >50% within 48 hours. If data were missing, we obtained the information by direct communication with attending doctors and other healthcare providers. Missing data were excluded from statistical analyses. All data were extracted by two physicians (XX and XX) independently, any disagreement was resolved by consensus or consulting a third researcher (XX).

### 2.4 Statistical analysis

All statistical analyses were performed using SPSS 23.0 software. Categorical variables were described as absolute numbers and percentages, and continuous variables were described using means and standard deviations. When the data were normally distributed, t test was used to compare the mean of the two samples; otherwise, the rank sum test was used. When comparing multiple groups, we used variance analysis or covariance analysis. χ 2 test or Fisher exact probability test were used for categorical data. The results of correlation analysis were presented as a scatter plot. We calculated Pearson correlation coefficients or partial correlation coefficients after controlling the confounding factors such as gender and age. P values less than 0.05 was considered statistically significant.

## 3. RESULTS

### 3.1 Characteristics of epidemiology and demography

Clinical data of 32 children with confirmed SARS-CoV-2 infection were collected in this study, with 20 (63%) patients from Chongqing and 3 (9%) patients from Shaanxi Province. The average age of the patients was nine years (SD=4.7), of which ten (31%) patients were under six years of age, 11 (34%) aged between six to 12 years and 11 (34%) aged between 12 to 18 years. 17 (53%) patients were male. Only 3 patients had underlying comorbidities: 1 had thrombocytopenia, 1 had Down’s syndrome and 1 had intestinal atresia. The vital signs on admission including average temperature, respiratory rate and heart rate were 36.7 °C (SD=0.5),23 bpm (SD=4) and 95 bpm (SD=12), respectively. 12 (38%) patients had a history of travel or living in the affected area and 29 (91%) patients with known cases among family members. All patients received antiviral treatment, of which interferon (94%) and lopinavir/ritonavir tablets (47%) were the most commonly used. (Table 1)

**Table 1.**
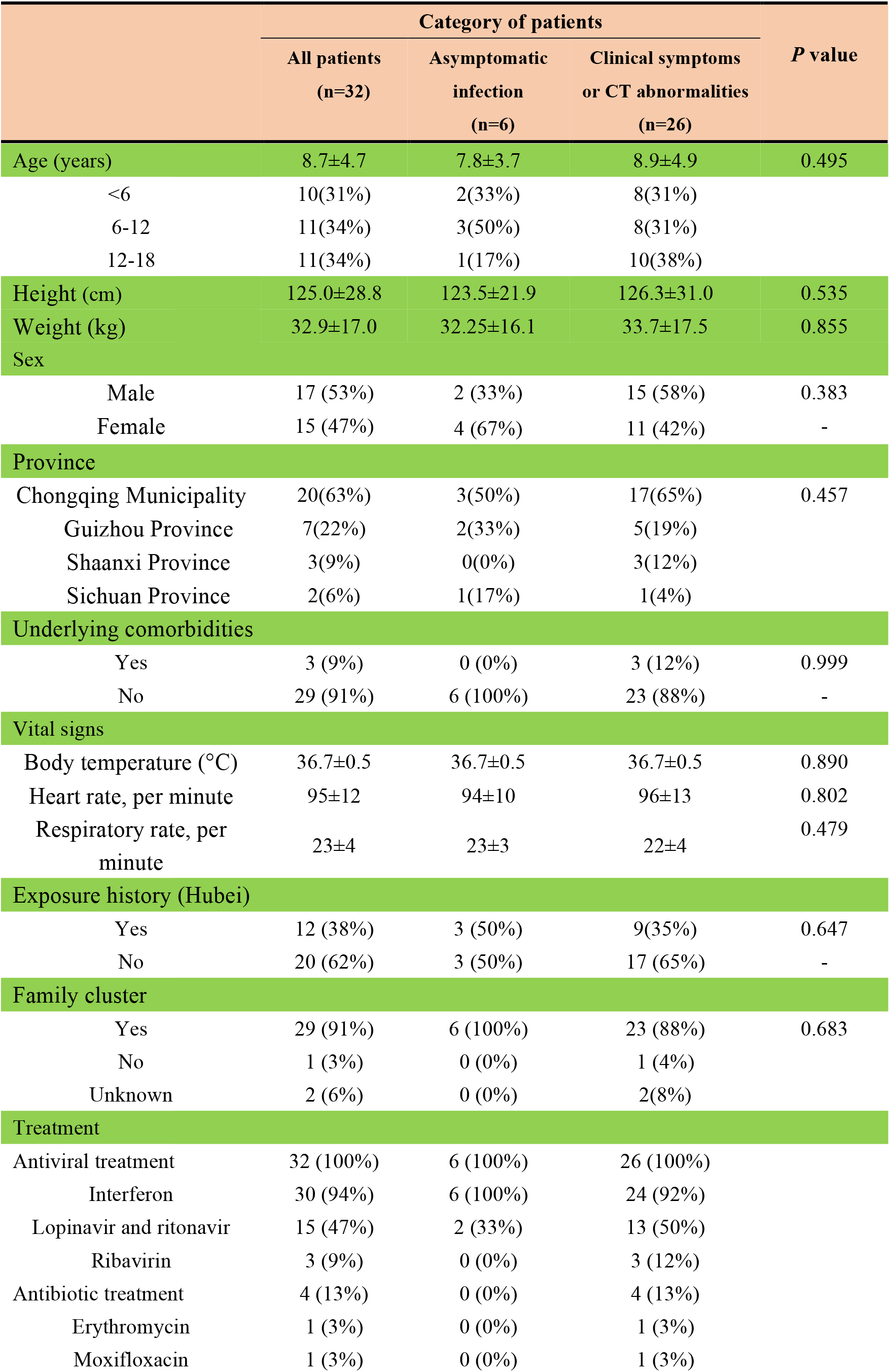
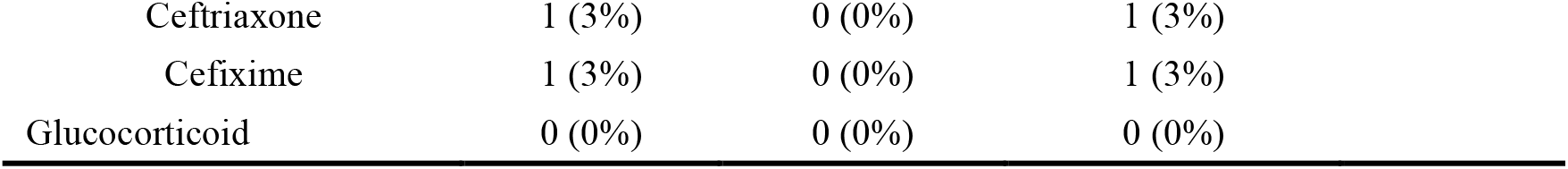
Baseline characteristics and exposure history of children infected with SARS-CoV-2.

|  | Category of patients |  |  | *P* value |
| --- | --- | --- | --- | --- |
|  | All patients<br>(n=32) | Asymptomatic<br>infection<br>(n=6) | Clinical symptoms<br>or CT abnormalities<br>(n=26) |  |
| Age (years) | 8.7±4.7 | 7.8±3.7 | 8.9±4.9 | 0.495 |
| <6 | 10(31%) | 2(33%) | 8(31%) |  |
| 6-12 | 11(34%) | 3(50%) | 8(31%) |  |
| 12-18 | 11(34%) | 1(17%) | 10(38%) |  |
| Height (cm) | 125.0±28.8 | 123.5±21.9 | 126.3±31.0 | 0.535 |
| Weight (kg) | 32.9±17.0 | 32.25±16.1 | 33.7±17.5 | 0.855 |
| Sex |  |  |  |  |
| Male | 17 (53%) | 2 (33%) | 15 (58%) | 0.383 |
| Female | 15 (47%) | 4 (67%) | 11 (42%) | - |
| Province |  |  |  |  |
| Chongqing Municipality | 20(63%) | 3(50%) | 17(65%) | 0.457 |
| Guizhou Province | 7(22%) | 2(33%) | 5(19%) |  |
| Shaanxi Province | 3(9%) | 0(0%) | 3(12%) |  |
| Sichuan Province | 2(6%) | 1(17%) | 1(4%) |  |
| Underlying comorbidities |  |  |  |  |
| Yes | 3 (9%) | 0 (0%) | 3 (12%) | 0.999 |
| No | 29 (91%) | 6 (100%) | 23 (88%) | - |
| Vital signs |  |  |  |  |
| Body temperature (°C) | 36.7±0.5 | 36.7±0.5 | 36.7±0.5 | 0.890 |
| Heart rate, per minute | 95±12 | 94±10 | 96±13 | 0.802 |
| Respiratory rate, per minute | 23±4 | 23±3 | 22±4 | 0.479 |
| Exposure history (Hubei) |  |  |  |  |
| Yes | 12 (38%) | 3 (50%) | 9(35%) | 0.647 |
| No | 20 (62%) | 3 (50%) | 17 (65%) | - |
| Family cluster |  |  |  |  |
| Yes | 29 (91%) | 6 (100%) | 23 (88%) | 0.683 |
| No | 1 (3%) | 0 (0%) | 1 (4%) |  |
| Unknown | 2 (6%) | 0 (0%) | 2(8%) |  |
| Treatment |  |  |  |  |
| Antiviral treatment | 32 (100%) | 6 (100%) | 26 (100%) |  |
| Interferon | 30 (94%) | 6 (100%) | 24 (92%) |  |
| Lopinavir and ritonavir | 15 (47%) | 2 (33%) | 13 (50%) |  |
| Ribavirin | 3 (9%) | 0 (0%) | 3 (12%) |  |
| Antibiotic treatment | 4 (13%) | 0 (0%) | 4 (13%) |  |
| Erythromycin | 1 (3%) | 0 (0%) | 1 (3%) |  |
| Moxifloxacin | 1 (3%) | 0 (0%) | 1 (3%) |  |

|  |  |  |  |
| --- | --- | --- | --- |
| Ceftriaxone | 1 (3%) | 0 (0%) | 1 (3%) |
| Cefixime | 1 (3%) | 0 (0%) | 1 (3%) |
| Glucocorticoid | 0 (0%) | 0 (0%) | 0 (0%) |

### 3.2 Clinical characteristics and radiological findings

Of the confirmed 32 patients, eleven children (34%) patients were asymptomatic with six (19%) asymptomatic patients had normal CT scan. At admission, the most frequently experienced symptoms were fever (38%) and cough (57%). According to CT scans, 19 (59%) patients showed abnormalities in the lungs (suggesting pneumonia), of whom three (9%) patients showed bilateral pneumonia, eight (25%) patients showed unilateral pneumonia and eight (25%) patients showed multiple mottling and ground-glass opacity. Five (16%) patients were found to have only CT abnormalities without clinical symptoms. (Table 2)

**Table 2.** Clinical characteristics of children infected with SARS-CoV-2 (n=32)

| **Clinical characteristics** |  |
| --- | --- |
| Fever | 8/21(38%) |
| Cough | 12/21(57%) |
| Fever and cough | 6/21(29%) |
| Stuffy nose | 3/21 (14%) |
| Runny nose | 5/21 (24%) |
| Sore throat | 3/21 (14%) |
| Headache | 2/21 (10%) |
| Shortness of breath | 0/21 (0%) |
| Shiver | 1/21 (5%) |
| Choking sensation in Chest | 0/21 (0%) |
| Fatigue | 2/21 (10%) |
| Asymptomatic | 11/32 (34%) |
| Asymptomatic and normal CT | 6/32 (19%) |
| Only abnormalities on CT | 5/32 (16%) |
| More than one symptom | 11/21 (52%) |
| **CT findings** |  |
| Abnormal | 19 (59%) |
| Unilateral pneumonia | 8 (25%) |
| Bilateral pneumonia | 3 (9%) |
| Multiple mottling or ground-glass opacity | 8 (25%) |

### 3.3 Laboratory findings

On admission, leucocytes were above the normal range in two (6%) patients and below the normal range in one (3%) patient, with an average number of total leucocytes being 6.1 x10^9^/L (SD=0.4). The average number of total leucocytes was 8.6 x10^9^/L (SD=1.6) in children with asymptomatic infection and 5.5 x10^9^/L (SD=0.3) in children with clinical symptoms and/or CT abnormalities, which is significantly lower than that of asymptomatic patients (P=0.011). Children with clinical symptoms and/or CT abnormalities were subdivided into three groups as children with CT abnormalities only, clinical symptoms only and both, where no significant difference in leucocyte counts was found. Neutrophils were above the normal range in eight (25%) patients, with an average value of 3.0 x10^9^/L(SD=0.4). The average counts of neutrophils was 4.9 x10^9^/L (SD=1.6) in children with asymptomatic infection and 2.5 x10^9^/L(SD=0.2)in children with clinical symptoms and/or CT abnormalities, which is significantly lower than that of children with asymptomatic infection (P=0.006). There were no significant differences between children with CT abnormalities only, children with clinical symptoms only and both in neutrophils counts. Lymphocytes were below the normal range only in two (6%) patients, while nine (28%) patients showed high lymphocytes counts above the normal range, with an average of 2.6 x10^9^/L (SD=0.2). No significant differences were found between asymptomatic group and children with symptoms and/or CT abnormalities. Platelet counts and hemoglobin concentration were basically within the normal range.

Most patients had normal liver function: only one (3%) patient had elevated aspartate aminotransferase (AST) and six (19%) patients had elevated alanine aminotransferase (ALT), with the maximum value of ALT to be 80U/L. There were no significant differences between children with asymptomatic infection and those with clinical symptoms and/or CT abnormalities. Albumin reduction was seen in only one patient. Eleven (37%) patients had increased LDH values. The bilirubin level in children who had CT abnormalities with or without clinical symptoms was lower than those with clinical symptoms only (P=0.022), with four (12%) cases with bilirubin levels above the normal range. For the coagulation function, APTT and PT were within the normal range, and D-dimer was above the normal range in six (24%) patients among 25 patients tested,in which the children who had CT abnormalities with or without clinical symptoms showed lower D-dimer than those with clinical symptoms only (P=0.031). As for bacterial co-infection, procalcitonin was below 0.5ug/L for all patients and three (12%) patients showed increased CRP values among the 26 patients tested. (Table 3)

**Table 3.**
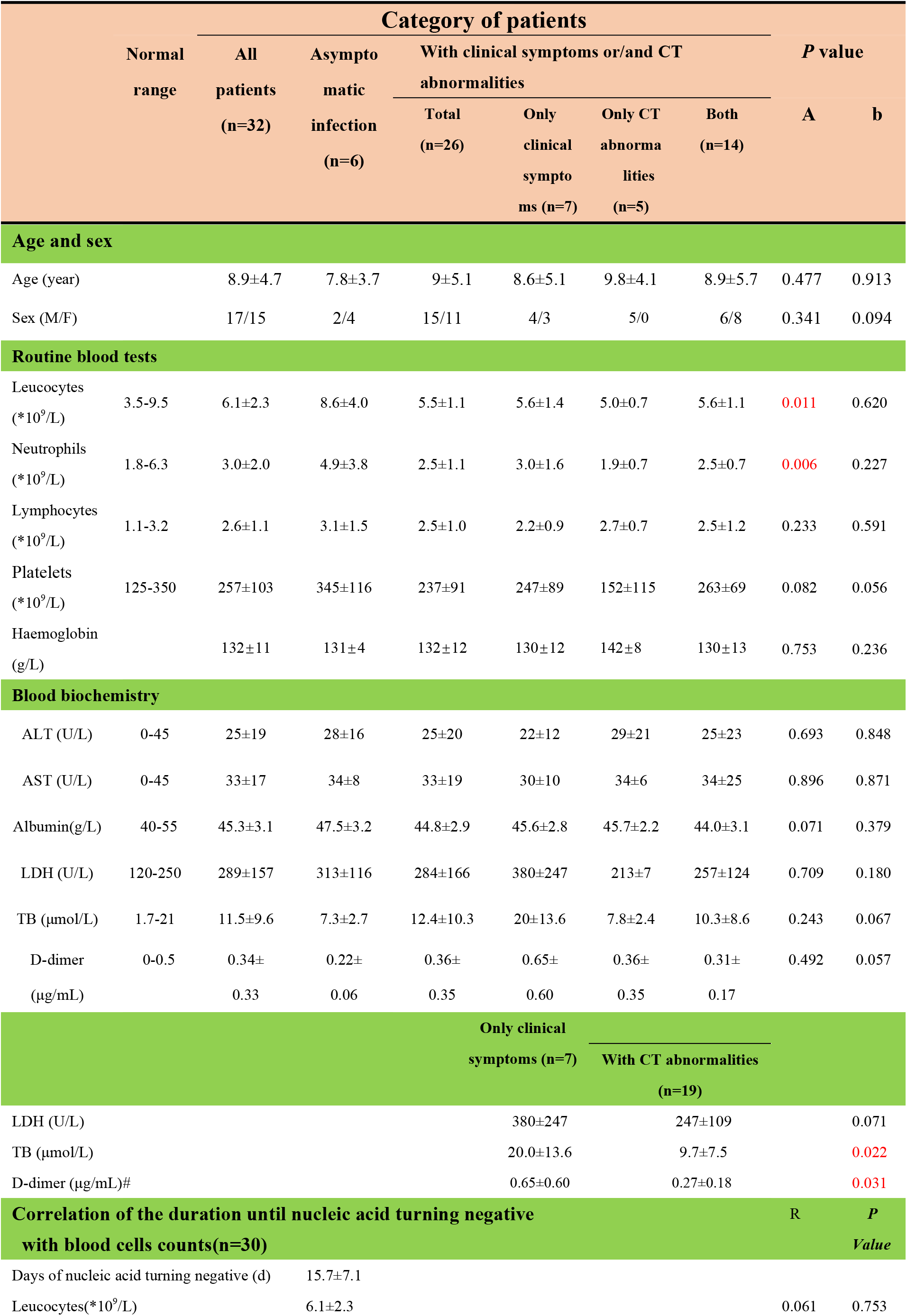
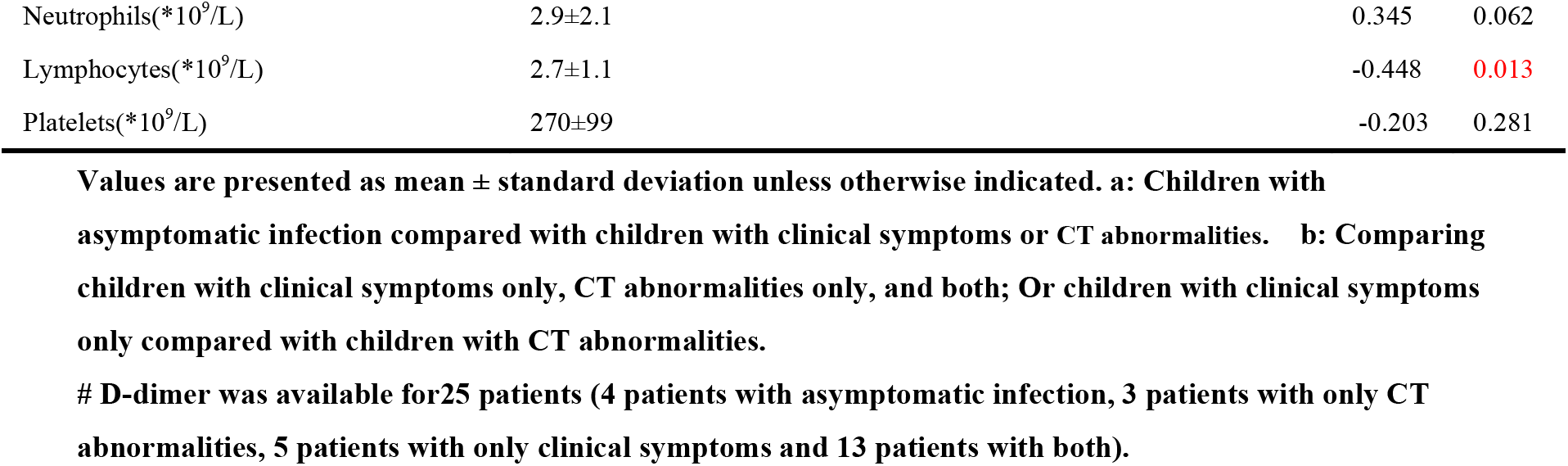
Laboratory results of children with SARS-CoV-2 infection.

### 3.4 Analysis of duration of positive nucleic acid and recurrence of positive results

#### 3.4.1 SARS-CoV-2 duration of positive nucleic acid

By the end of February 29, all 32 patients had been discharged and no severe cases or deaths occurred. Among the 32 children except one lost some re-tested data who had tested positive for SARS-CoV-2 nucleic acid, by 14-days 52% (16/31) had a negative nucleic acid test. This was 33% (2/6) and 56% (14/25) for asymptomatic patients and symptomatic patients, respectively, which was not statistically different. The mean time from the first positive or onset of symptoms until the first negative nucleic acid test was 15.4 (SD=7.2) days in 31 patients. No significant difference was identified between children with asymptomatic infection (15.0 days, SD=6.7) and children with clinical symptoms (15.5 days, SD=7.4). In addition, 14 parents, of the enrolled children, who were themselves infected showed a mean duration of 22.7 (SD=10.0) days until first negative nucleic acid test, slightly longer than that in children (P=0.07) (Table 3). The mean duration of until the first negative nucleic acid test was 12.4 days, 18.6 days and 14.9 days in patients under 6 years, 6 to 12 years and 12 to 18 years, respectively, with no statistically significant difference between these three age-groups (Table 4).

**Table 4.** Time until SARS-Cov-2 nucleic acid turning negative in the children and their parents.

|  | Category of patients |  |  | *P*<br>value |
| --- | --- | --- | --- | --- |
|  | All patients<br>(n=32) | Asymptomatic<br>infection (n=6) | Clinical symptoms or<br>CT abnormalities (n=26) |  |
| Nucleic acid turned negative within 14 days |  |  |  |  |
| Yes | 16(52%) | 2 (33%) | 14 (56%) | 0.394 |
| NO | 15 (48%) | 4 (67%) | 11 (44%) | - |
| Unknown | 1 | 0 | 1 | - |
| Days of nucleic acid turning negative (d) |  |  |  |  |
|  | 15.4±7.2 | 15.0±6.7 | 15.5±7.4 | 0.853 |
| Children | 17.6±7.9 |  |  | 0.070 |
| *Parents (n=17) | 22.7±10.0 |  |  |  |

\*Seventeen parents of the participating children who were infected themselves

#### 3.4.2 Recurrence of positive SARS-CoV-2 nucleic acid results

Recurrence of positive SARS-CoV-2 nucleic acid test results were found in four (29%) children five to 30 days after the first negative nucleic acid test among 14 children who had subsequent testing after a negative result, of whom one patient was from the asymptomatic group and another three were from children with clinical symptoms and/or CT abnormalities. No significant difference between these two groups was identified (Table 5). And anal swab was tested on 6 children on follow up after discharge and quarantine 14 days, of which 5 tested positive (Table 6).

**Table 5.** Time of SARS-Cov-2 nucleic acid turning negative according to age.

| **Age(year)** | **Time of nucleic acid turning**<br>**negative (d)** | ***P* value** |
| --- | --- | --- |
| <6 | 11.5±3.7 | 0.360 |
| 6-12 | 18.6±9.4 |  |
| 12-18 | 14.9±5.9 |  |

**Table 6.**
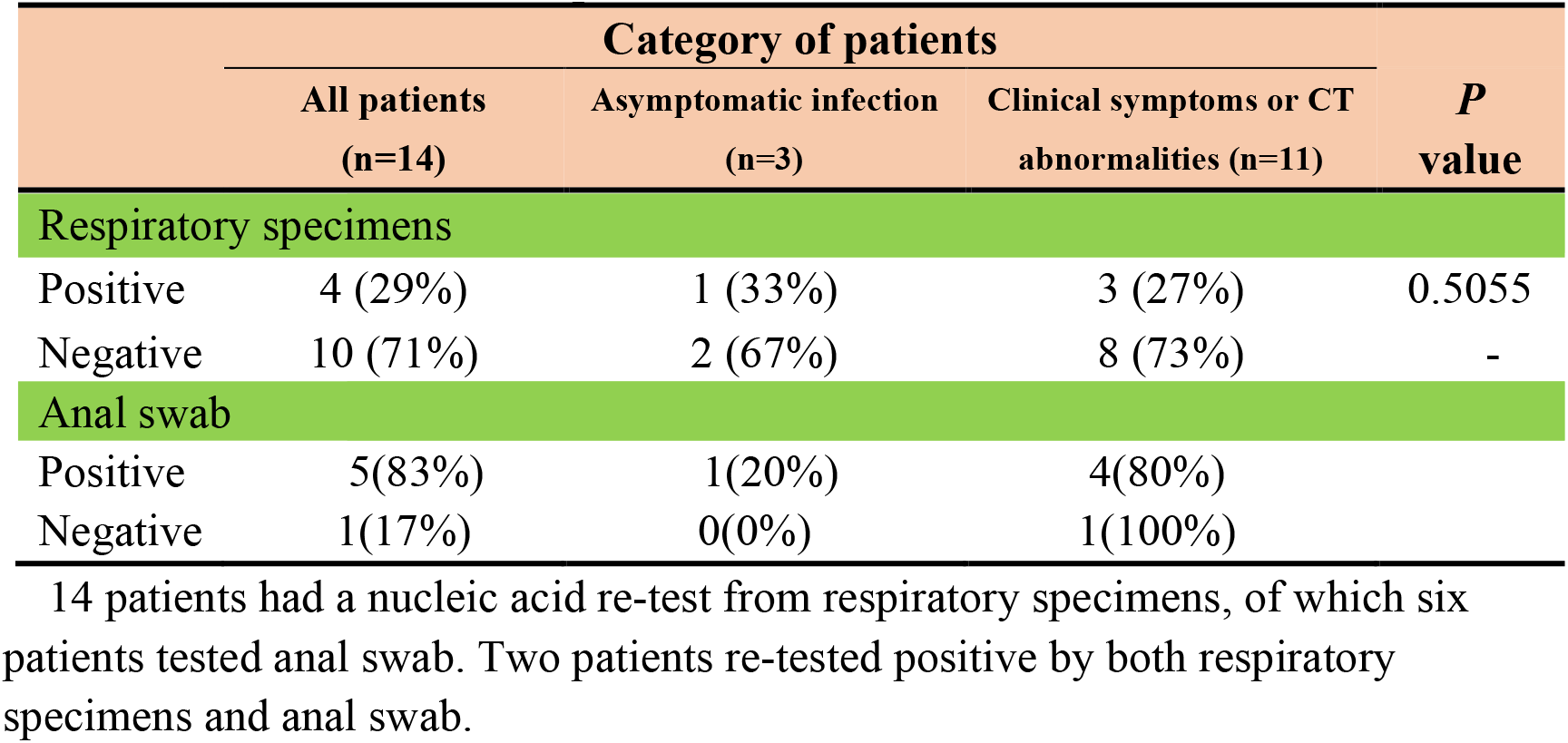
Recurrence of positive SARS-CoV-2 nucleic acid in children.

### 3.5 Correlation of the duration until first negative nucleic acid test with blood cells counts

Among the 32 cases, one patient failed to obtain a complete nucleic acid test result. Another patient showed significantly increased leucocytes after reexamination during hospitalization, which increased from 6.29 x10^9^/L to 14.34 x10^9^/L, indicating the presence of co-infection that affected the real situation of routine blood tests, thus these two patients were excluded from data analysis. We found a significant negative correlation between the lymphocyte counts and the time until the first negative nucleic acid, after adjusting for age, gender and length of stay (LOS). In other words, the more lymphocytes, the shorter the time of positive viral nucleic acid, which indirectly reflects the faster virus clearance (Table 3).

## DISCUSSION

The follow-up study is the first time to clarify the clinical characteristics, the time of nucleic acid turning negative and its correlation with lymphocyte counts between children with asymptomatic SARS-CoV-2 infection and those with clinical symptoms. About 38% of the children participating in our study had fever and 57% had cough at the time of admission to hospital. For most children, cough was the first symptom. In CT imaging, pulmonary changes in children were less severe than those in adults, with lower incidence of further progress to severe pneumonia.

Evidence on COVID-19 from adults suggested that at the time of admission, the leucocyte counts was in normal range or decreased in 70% to 76% of patients, while the absolute value of lymphocytes was significantly below the normal range in 35% to 63% of patients ^[12-13]^. In contrast, the results of our study showed that the absolute value of lymphocytes in children with SARS-CoV-2 was mostly normal with only 6% below the normal range. However, the published expert consensus of COVID-19 for children recommends that clinicians should take decreased lymphocyte counts as one of the diagnostic criteria for suspected cases ^[23-24]^. Based on the analysis of these 32 patients in our study, we suggested that using decreased lymphocyte counts for the diagnosis of COVID-19 in children may lead to missed diagnoses of suspected cases and cannot be recommended for diagnosis for children. In addition, our study also found that the total number of white blood cells and absolute neutrophils in children with clinical symptoms and/or CT abnormalities were lower than those with asymptomatic infection, while the lymphocyte counts showed no reduction. These data suggested that children with SARS-CoV-2 infection may have milder inflammatory response considering that neutrophils were closely related to inflammation, which may be one of the causes of mild illness in children.

Studies from adults showed 98% of patients had decreased albumin. Liver function in children are less affected by COVID-19 compared with adults ^[12]^. Increased D-dimer was found in 24% of patients who were all symptomatic while asymptomatic patients showed no abnormalities. The level of D-dimer and total bilirubin in children with only clinical symptoms was higher than those who had CT abnormalities with or without clinical symptoms, indicating that D-dimer was kept lower in children with pulmonary imaging abnormalities and that the virus may partially affect the synthesis of bilirubin and D-dimer, of which the significance and mechanism need to be further studied.

As for treatment measures, all included children received antiviral treatment (mostly interferon and lopinavir/ritonavir) and antibiotics, antifungal agents, glucocorticoid therapy were not used. Previous studies showed that the majority of adults with COVID-19 were given antiviral treatment (76%) and antibiotic treatment (71%), and some patients needed antifungal agents (15%) for co-infection control. In some severe cases, oxygen therapy, IVIG and glucocorticoid was need as adjuvant treatment in 76%, 27% and 19% of the patients, respectively ^[12]^.

Previous studies have shown that 26% to 32% of patients in adults were admitted to the intensive care unit (ICU) because of their higher oxygen requirement due to hypoxemia. Nosocomial infection was presented in 41% of adults with high risk of death (4.3% to 15%) ^[12-14]^. However, in our study, all patients recovered after treatment and no one died or was admitted to the pediatric intensive care unit (ICU) for any other oxygen support. The clinical outcome of COVID-19 in children seems to be significantly better than that in adults.

The mean duration of a positive nucleic acid test, until the first negative result, in children was 15 days, with no significant difference between asymptomatic infected children and children with symptoms or CT abnormalities. Neither was there any statistical difference between that of children and their parents, which was consistent with recent study ^[25]^. which was consistent with the previously reported 14-day incubation period. These results underline that virus replication occurs for a similar period in children with asymptomatic infection, which implies that children with asymptomatic infection should be quarantined for the same duration as symptomatic patients infected with SARS-CoV-2.

No significant difference in the duration of a positive nucleic acid test, until the first negative result was identified between children of different ages, but it seemed that the nucleic acid test became negative faster in children under six years of age, which may be associated with faster virus clearance. However, further research is needed to address this. Recurrence of positive SARS-CoV-2 nucleic acid test results were found in four children five to 30 days after the first negative nucleic acid test, all of them however showed improved clinical symptoms, as with adults ^[26]^. Recurrence of positive SARS-CoV-2 nucleic acid test results is not necessarily related to recurrent infection and also needs further investigation. In addition, we found a significant negative correlation between the lymphocyte count and the time until the first negative nucleic acid. This result implies that lymphocytes have an impact on the inhibition of SARS-CoV-2 replication, which may partially explain the reason why most children infected with SARS-CoV-2 present with only mild illness.

## Conclusions

The clinical characteristics, radiological and laboratory findings, and clinical outcomes of children infected with SARS-CoV-2 are essentially different from the situation in adults. Besides, there were also differences in laboratory findings between patients with asymptomatic infection and patients with symptoms or CT abnormalities. The clinical significance and mechanism behind the negative correlation between the number of lymphocytes and the time of SARS-CoV-2 nucleic acid turning negative need to be further studied. This study has limitations including the small sample size and partially missing of data in laboratory test, however, our study can be considered the best evidence yet for management of children infected with SARS-CoV-2 considering the current situation of public health emergency. Therefore, prospective and large-sample studies are needed for further research on children infected with SARS-CoV-2 to provide higher-quality and more credible evidence for clinical practice.

## Data Availability

All data generated or used during the study appear in the submitted article.

## Acknowledgement

We are very grateful to all the medical, nursing and supportive staff of the Chongqing Public Health Medical Center, Yongchuan Hospital of Chongqing Medical University, Three Gorges Hospital, Third Affiliated Hospital of Zunyi Medical University, Xi’an Children’s Hospital, Tongchuan Mining Bureau Central Hospital, the People’s Hospital of Xiushan County, the People’s Hospital of Deyang City, Nanchong Central Hospital, Suining Central Hospital, the People’s Hospital of Fengdu County, the People’s Hospital of Kaizhou District, the People’s Hospital of Hechuan District, the People’s Hospital of Fengjie County, Guizhou Provincial People’s Hospital, for their dedication in looking after the patients.

## Footnote

### Ethical Statement

The authors are accountable for all aspects of the work in ensuring that questions related to the accuracy or integrity of any part of the work are appropriately investigated and resolved. This study strictly complied with the ethical requirements of biomedical research issued by the relevant international and national organizations and was approved by the Ethics Committee of Children’s Hospital of Chongqing Medical University (No. 2020-002). Given the urgency in policy and clinical decision-making for COVID-19 and difficulty of confirming information for most patients, informed consent was exempted for all patients after discussion and approval of the above-mentioned committee.

